# Vitamin D status associates with skeletal muscle loss after anterior cruciate ligament reconstruction

**DOI:** 10.1101/2022.11.02.22281843

**Authors:** Jean L. Fry, Angelique N. Moore, Christine M. Latham, Katherine L. Thompson, Nicholas T. Thomas, Brooke D. Lancaster, Christopher S. Fry, Kelsey A. Reeves, Brian Noehren

## Abstract

**Objective:** We evaluated associations between vitamin D status and skeletal muscle, strength, and bone mineral density (BMD) outcomes after ACL reconstruction (ACLR) in an observational study.

**Methods:** Serum measures included 25-hydroxyvitamin D (25(OH)D; free and total), vitamin D binding protein (DBP), and 1,25-dihydroxy vitamin D (1,25(OH)_2_D) at baseline, 1 week, 4 months, and 6 months post-ACLR. Vastus lateralis biopsies were collected from the healthy and ACL-injured limb of 21 young, healthy participants (62% female; 17.8 [3.2] yr, BMI: 26.0 [3.5] kg/m^2^) during ACLR and the injured limb only at 1 week and 4 month follow ups. RNA and protein were isolated from biopsies and assessed for vitamin D receptor [*VDR*], and vitamin D-activating enzymes. Quadriceps fiber cross-sectional area (CSA) was determined with immunohistochemistry. BMD of femur and tibia were determined at baseline and 6 months post-ACLR; strength was assessed with an isokinetic dynamometer.

**Results:** 1,25(OH)_2_D decreased from baseline to one week after ACLR (21.6 [7.9] vs. 13.8 [5.5] pg/mL; p<0.0001). *VDR* and 25-hydroxylase transcript abundance and VDR and DBP proteins were elevated one week after ACLR compared with baseline (FDR<0.05; p<0.05). Participants with an average total 25(OH)D <30 ng/mL showed significant decreases in CSA 1 week and 4 months after ACLR (p<0.01; p=0.041 for time x D status interaction), whereas those with total 25(OH)D ≥30ng/mL showed no significant differences (p>0.05 for all comparisons). BMD and strength measures were lower at follow up but did not associate with vitamin D status.

**Conclusion:** ACLR promotes vitamin D pathways in the quadriceps and low status is associated with loss of skeletal muscle both 1 week and 4 months after ACLR.

**Summary Box:**

- **What is already known on this topic** – Quadriceps muscle atrophy, strength loss, and reduced bone mineral density persist for many years after ACL tear and reconstruction (ACLR) leading to poorer function and long term knee health outcomes. Circulating 25-hydroxyvitamin D concentrations ≥30ng/mL (75nmol/L) have been associated with reduced risk of stress fracture and injury and greater strength, but it is not known how vitamin D status, which is easily modified with supplementation, may affect ACLR outcomes.
- **What this study adds** – Our work shows that ACLR surgery reduces biologically active vitamin D in circulation and promotes vitamin D receptor and activating enzyme expression in skeletal muscle one week after surgery. Circulating concentrations of 25(OH)D <30 ng/mL associate with greater loss of quadriceps fiber CSA both one week and 4 months after ACLR.
- **How this study might affect research, practice or policy** – Results suggest that correcting vitamin D status prior to ACLR may support retention of skeletal muscle size in recovery, which should be tested in a randomized clinical trial to begin to establish vitamin D cut points optimizing recovery from ACL tear and reconstruction.

## Introduction

Anterior cruciate ligament injury and reconstruction (ACLR) promotes long term deficits in lower limb structure and function. Even several years after ACLR, 20-40% reductions in quadriceps strength remain [1], which associates with poorer function continuing for at least 3 years after ACLR [2, 3]. Bone mineral density (BMD) in the proximal tibia and distal femur significantly decrease within 6 months after ACLR [4, 5], and patients show significant reductions in distal femur BMD that persist for at least 2 years [5]. These strength and BMD deficits substantially increase risk for development of knee osteoarthritis [6, 7]. Many potential causes have been evaluated, but clinicians still need interventions that reliably improve patient outcomes [8].

Little is known about how vitamin D status affects ACL outcomes. Vitamin D, an endogenously synthesized steroid hormone and dietary component, is unequivocally essential for proper bone mineralization in youth [9]. Vitamin D functions primarily through hormone activity by binding to the its nuclear receptor (VDR) and regulating approximately 2000 human genes [10]. Circulating 25-hydroxyvitamin D (25(OH)D) concentrations of ≥30ng/mL (75nmol/L), which is higher than minimum cut points established for deficiency [9], have been associated with reduced risk of stress fractures and sports injury [11, 12], greater grip strength [13], and several other health outcomes [14-16]. Participants with higher vitamin D status show better muscle strength and recovery [17, 18] and supplementation may improve lower body strength in athletes [19]. Studies in preclinical models show that VDR overexpression causes muscle hypertrophy [20] and knockdown of VDR induces muscle atrophy [21]. Chemical injury in rodent skeletal muscle promotes expression of VDR and CYP27B1 (a vitamin D activating enzyme) [22]. Despite the apparent benefits of vitamin D for skeletal muscle health, the VDR is barely detectable in healthy, mature human skeletal muscle tissue [23, 24].

Anterior cruciate ligament (ACL) tears and reconstruction (ACLR) result in long-term protracted muscle weakness [25], and effective therapies to fully prevent muscle and strength loss related to ACL tear and ACLR remain elusive. Here we sought to characterize vitamin D-associated activity in quadriceps muscle after ACLR and determine how vitamin D status associates with skeletal muscle size, bone mineral density and strength outcomes. We hypothesized that ACL reconstruction would induce expression vitamin D-related pathways and that optimal vitamin D status (25(OH)D ≥ 30ng/mL) would associate with better retention of strength, power and size of skeletal muscle and less loss of BMD in the injured limb. Our primary outcome was mean quadriceps muscle fiber cross sectional area.

## Participants and Methods

All study protocols were approved by the University of Kentucky (UK) Institutional Review Board. All participants provided written and oral consent prior to data collection or parental consent and child assent, where applicable. All participants (n=21) were recruited after an ACL injury and before ACLR surgery, were between 15-29 years of age (median 17 years), underwent bone-patellar-bone graft ACLR conducted at the UK Orthopedic Surgery & Sports Medicine practice in Lexington, KY. Thereafter, participants completed a progressive rehabilitation program according to previously published guidelines at the UK’s Physical Therapy Department [26, 27]. Complete methods are provided in supplement 1. Participant characteristics are detailed in Table 1. Fig. 1 provides an overview of sample and data collection.

**Table 1.**
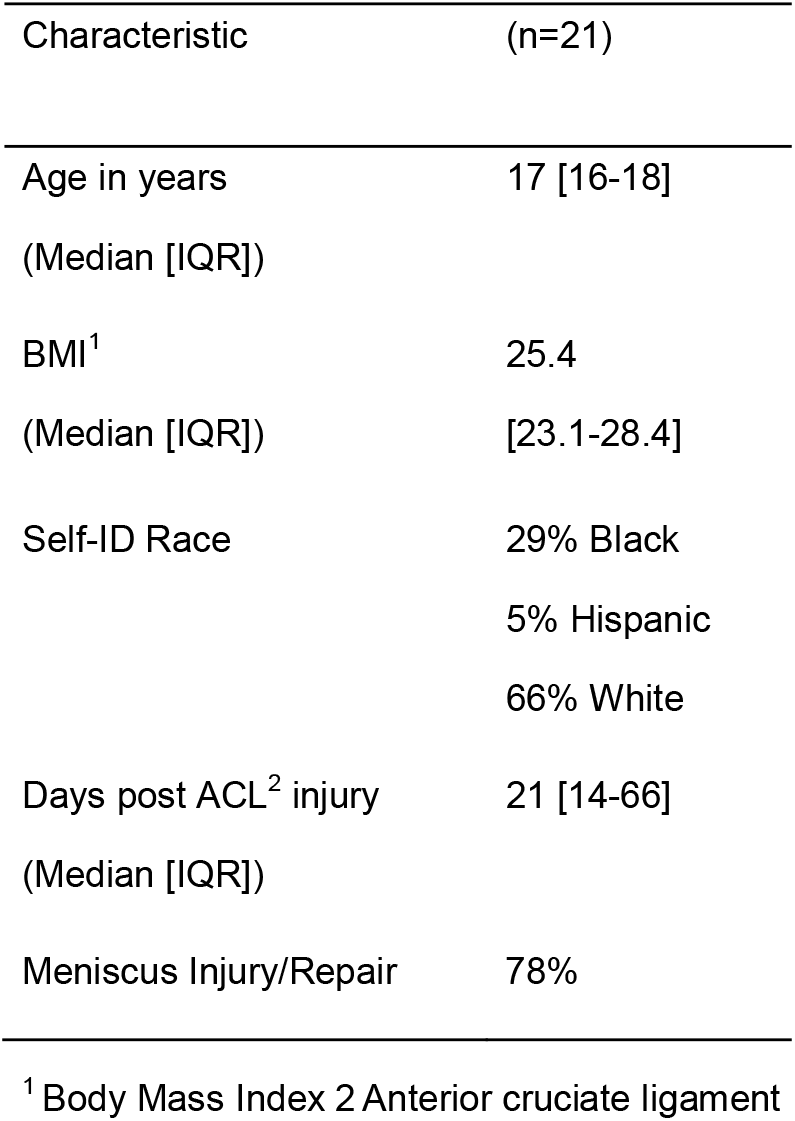
Participant demographics at study baseline.

| Characteristic | (n=21) |
| --- | --- |
| Age in years<br>(Median [IQR]) | 17 [16-18] |
| BMI<sup>1</sup><br>(Median [IQR]) | 25.4<br>[23.1-28.4] |
| Self-ID Race | 29% Black<br>5% Hispanic<br>66% White |
| Days post ACL<sup>2</sup> injury<br>(Median [IQR]) | 21 [14-66] |
| Meniscus Injury/Repair | 78% |

<sup>1</sup> Body Mass Index 2 Anterior cruciate ligament

**Table 1.** Participant demographics and baseline characteristics

| Characteristic | (n=21) |
| --- | --- |
| Age in years |  |
| (Median [IQR]) | 17 [16-18] |
| BMI<sup>1</sup> | 25.4 |
| (Median [IQR]) | [23.1-28.4] |
| Self-ID Race | 29% Black |
|  | 5% Hispanic |
|  | 66% White |
| Days post ACL<sup>2</sup> injury |  |
| (Median [IQR]) | 21 [14-66] |
| Meniscus Injury/Repair | 78% |

<sup>1</sup> Body Mass Index
<sup>2</sup> Anterior cruciate ligament

**Fig. 1.**
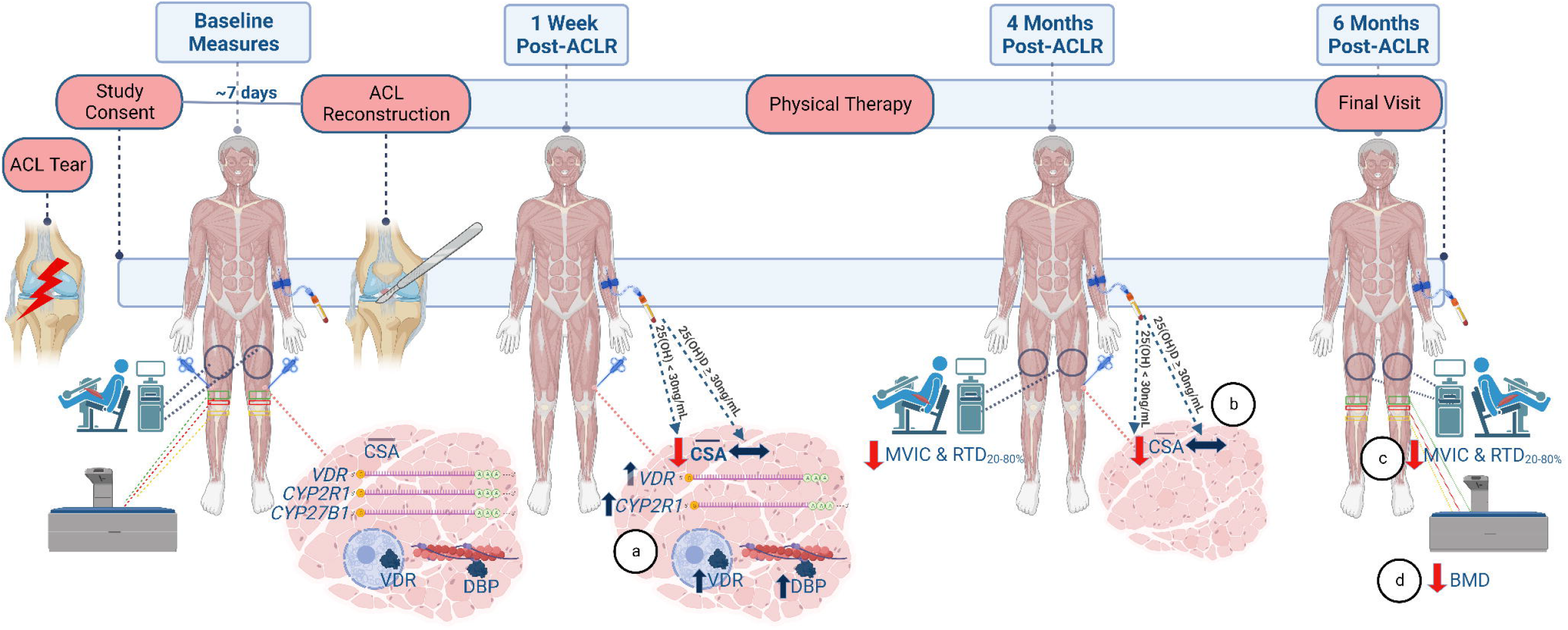
Study timeline and primary findings. 1a. The vitamin D receptor (VDR) and vitamin-D activating enzyme CYP2R1 were significantly increased at the transcript level one week after anterior cruciate ligament reconstruction (ACLR) when compared with baseline in the injured limb. VDR and vitamin D binding protein (DBP) protein were also increased at this time point. Participants with 25-hydroxyvitamin D (25(OH)D) <30ng/mL showed significant decreases in mean quadriceps fiber cross sectional area (CSA) one week after ACLR when compared with the injured limb at baseline; however no CSA changes were seen in those with 25(OH)D ≥30ng/mL. 1b. Participants with 25-hydroxyvitamin D (25(OH)D) <30ng/mL showed significant decreases in mean quadriceps fiber CSA 4 months after ACLR when compared with the injured limb at baseline. 1c. Maximal voluntary isometric contraction (MVIC) and rate of torque development (RTD_20-80%_) were significantly decreased among participant 6 months after ACLR, but there were no differences between vitamin D status groups. 1d. Bone mineral density in the femur and tibia, as measured by dual energy x-ray absorptiometry were significantly decreased among participants 6 months after ACLR, but there were no differences between vitamin D status groups.

### Patient and public involvement

Our ACL patients’ and participants’ demonstrated loss of muscle size and strength and bone mineral density after ACL tear reconstruction have inspired the research aims. Our group is committed to investigating ways to improve recovery from ACL and promote optimal long term function for these patients.

### Circulating Biomarkers

Analyzed serum samples were collected prior to ACLR, and at 1-week, 4 month, and 6 month follow up visits. Study vitamin D status was defined as the mean 25(OH)D over these 4 time points. Mayo Clinic Laboratories assessed 25(OH)D using gold standard LC-MS/MS methodology. An “optimization” cut point was established at 30 ng/mL (75 nmol/L) based on prior literature indicating optimal health outcomes at this concentration [11, 13, 15, 28]. ELISA was used to assess 1,25-dihydroxy vitamin D (1,25(OH)_2_D; Biovendor R&D RIS024R and RIS021R), free 25(OH)D (Biovendor R&D KAPF1991) and vitamin D binding protein (DBP; R&D Systems DY008B and DY3778B-05) according to the manufacturer’s instructions.

### Muscle Biopsies

Biopsies were taken from the vastus lateralis on the injured limb and contralateral healthy limb (control) at the time of ACLR and from the injured limb only 1 week and 4 months after ACLR for immunohistochemistry (IHC) analysis and protein/gene expression analyses [29]. The sample was divided and flash frozen for RNA/protein and for IHC mounted in tragacanth.

### Western Blot

Western blots from muscle biopsies were used to compare VDR and DBP protein before and after ACLR in the injured limb using the healthy limb as a control. Following homogenization, protein concentration was determined with the Bradford assay (Smartspec Plus spectrophotometer; Bio-Rad, Hercules, CA, USA) to enable us to load 50µg protein in each well. Samples were loaded on to stain-free gels with mouse kidney lysate (VDR positive control) and HELA lysate (loading control). Protein was transferred to a PVDF membrane and probed with VDR antibodies (Abcam ab109234; 1:1000), then stripped and blocked before incubating in DBP antibodies (0.25µg/mL; R&D Systems DY3778B-05 detection antibody). All blots were analyzed in ImageLab (Bio-Rad) by creating a multi-channel image with total protein coupled to the chemiluminescent channel. For each participant, all of their samples were loaded on the same gel.

### RNA isolation and sequencing (RNA-seq)

RNA was isolated from muscle homogenates in accordance with manufacturer guidelines (Direct-zol RNA Miniprep Kit, Zymo). RNA content, purity, and integrity was quantified using the 2100 Bioanalyzer (Agilent) (RIN > 8.5) and the NanoDrop 2000 (Thermo Fisher) at the University of Kentucky Genomics Core. Six hundred nanograms of total RNA was sent to Novogene Corporation (Chula Vista, CA) for library construction and sequencing on an Illumina HiSeq 4000 system using a paired-end 150 bp dual-indexing protocol. Raw FASTQ files underwent pre-alignment quality control, then were aligned using STAR aligner (reference genome: GRCh38/hg38) using Partek Flow, and analyzed to obtain differential gene expression using DESeq2 (minimum read cutoff of 50). Differential gene expression was calculated comparing ACL-injured samples collected during surgery and 1 week post-ACLR and comparing healthy limb samples collected during surgery and 1 week post-ACLR. Raw p-values were adjusted for multiple testing using the Benjamini-Hochberg false discovery rate (FDR) step-up method. Pathway over-representation analysis was performed using g:Profiler [30] with non-ordered query and up-or-downregulated genes with FDR < 0.05. RNA-sequencing data are deposited in Gene Expression Omnibus: GSE211681. Transcript abundance of vitamin D activating pathway regulators (vitamin D receptor [*VDR*], and Cytochrome P450 2R1 [*CYP2R1*] and Family 27 Subfamily B Member 1 [*CYP27B1*]) are reported here; the complete data set is available (GSE211681).

### IHC Analysis

Seven µm thick sections were rehydrated in PBS then incubated overnight in a rabbit anti-laminin primary antibody (Sigma L9393, diluted 1:100 in PBS). Slides were then washed and incubated in AlexaFluor 555 goat anti-rabbit secondary antibody (Invitrogen A21429, diluted 1:250 in PBS) for 2 hours, mounted with Vectashield mounting media (Vector Laboratories, cat# H-1000) and imaged on a Ziess AxioImager M2 upright fluorescent microscope. MyoVision, an automated image analysis software, was used to obtain resulting cross-sectional area data as previously described [31].

### Strength Outcomes

Participants’ weight-normalized maximum voluntary isometric contraction (MVIC, i.e. peak torque) and the mean slope of the torque-time curve between 20% and 80% of the first 200 milliseconds from muscle contraction onset (RTD_20-80%_) were evaluated before ACLR and at 4 and 6 month follow ups using previously reported protocols [32]. Participants completed MVIC and RTD_20-80%_ testing in both limbs using a Biodex 4 isokinetic dynamometer (Biodex Medical Systems Inc., Shirley, NY, USA). Results were analyzed with custom MATLAB code as previously described [32].

### Bone Density Measures

Bone mineral density (BMD) was assessed with dual energy x-ray absorptiometry (DXA) scans (Lunar iDXA, GE Healthcare) and were completed at study baseline and at the 6 month follow up in both the injured and healthy limbs. We utilized a validated protocol for determining BMD in the femur and tibia [33] in three regions, which are outlined in supplement 1. The DXA enCORE software platform automatically calculated bone mineral density.

### Statistical Analyses

For all outcomes, statistical significance was set at *p* < .05, using two-sided tests and using adjusted *p*-values where appropriate. All continuous measures were summarized with descriptive statistics, and distributions were tested for normality. Efforts were made to align with the BJSM checklist for statistical assessment of medical papers and we justify deviations were relevant [34]. Circulating measures taken at 4 time points were analyzed using a mixed-effects repeated measures analysis using the Tukey post-hoc test to adjust for multiple comparisons (n=21). Results of western blots were analyzed using paired t-tests utilizing data from the injured limb at baseline and the injured limb one week after surgery (n=18). To assess relationships between vitamin D status and outcomes (CSA, BMD, and Biodex measures), study 25(OH)D were redefined as a dichotomous variables based on the a priori optimization cut-off value of ≥30□ng/mL to determine high (n=8) and low status groups (n13). Analyzing vitamin D as a dichotomous variable with a cut point is 30ng/mL is common [13, 35, 36]. Since CSA, DXA, Biodex measures included multiple observations taken from the same subject over the injured/non-injured legs and across multiple visits, a full-factorial repeated-measures ANOVA was performed, first analyzing overall differences across the various treatment groups (time point/leg and vitamin D status). Likelihood ratio testing and Akaike Information Criterion (AIC) were used to select an appropriate covariance structure (here, compound symmetry covariance). A Kenward-Roger adjustment was used, as appropriate, to correct for negative bias in the standard errors and degrees of freedom calculations induced by small samples. For each relevant pairwise comparison, estimated differences of means (calculated as Group 1 - Group 2) and the associated standard errors are adjusted for baseline (V2) non-injured leg value. All analyses were completed in SAS 9.4 (SAS Institute Inc.; Cary, NC, USA).

## Results

### Circulating Biomarkers

Compared with baseline, 1-week post-ACLR circulating 1,25(OH)_2_D was substantially reduced (21.58 ± 7.88 pg/mL vs. 13.84 ± 5.46 pg/mL; p<0.0001 for baseline vs. w-week; p<0.01 main effect for time). Compared with baseline concentrations, DBP was significantly reduced at 4 and 6 months (233.17 ± 103.43 µg/mL vs. 200.50 ± 99.22 µg/mL and 192.04 ± 73.61 µg/mL; p<0.001 for main effect). Circulating total and free 25(OH)D did not change significantly throughout the study. Results of circulating biomarkers are presented in Fig. 2 and complete time course data for all participants is included in supplemental table S1 and by vitamin D status group in supplemental table S2.

**Fig. 2.**
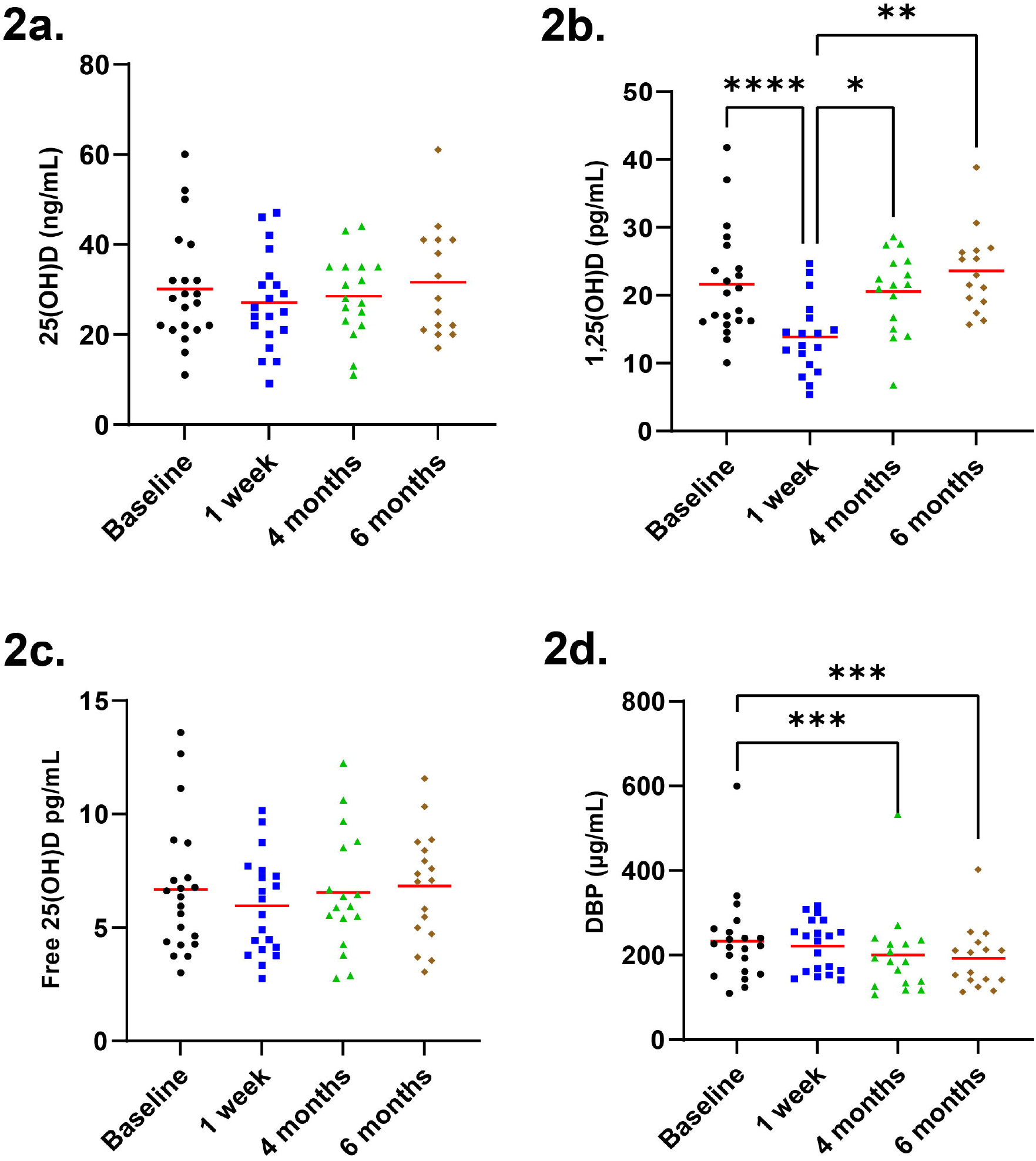
Circulating vitamin D metabolites before and after ACL reconstruction (ACLR; n=21). 2a. 25-hydroxy vitamin D (25(OH)D; status indicator) unchanged throughout the study (p=0.360). 2b. 1,25-dihydroxy vitamin D (1,25(OH)_2_D; active form) is significantly reduced after ACLR (p<0.001). 2c. Free 25(OH)D unchanged throughout the study (p=0.433). 2d. Vitamin D Binding Protein (DBP) lower at 4 and 6 month follow up when compared with baseline and 1 week post ACLR (p=<.0001). One-way repeated measures ANOVA; results of host-hoc tests on graph: * p-value <0.05, ** p-value <0.01, **** p-value <0.0001.

### Vitamin D Pathways are Elevated in Skeletal Muscle After ACLR

Concentrations of VDR protein in the quadriceps increased in the injured limb after ACLR (0.2 ± 0.2 arbitrary units (AU) vs. 0.7 ± 0.7 AU; p=0.008). DBP content is also modestly increased in the injured limb after ACLR (0.66 ± 0.53 AU vs. 0.81 ± 0.32; p=0.047). Complete data is presented in supplemental tables S3 and S4 and data from the injured limb is represented in Fig. 3a and 3b. RNA-seq data and analysis showed elevations in VDR (FDR<0.05), CYP2R1 (FDR<0.05) when comparing the 1-week measure to both baseline injured and healthy limbs. There were no significant changes in CYP27B1. Transcript abundance data from the RNA-seq data set are presented in supplemental table 2 and Fig. 3c and 3d.

**Fig. 3.**
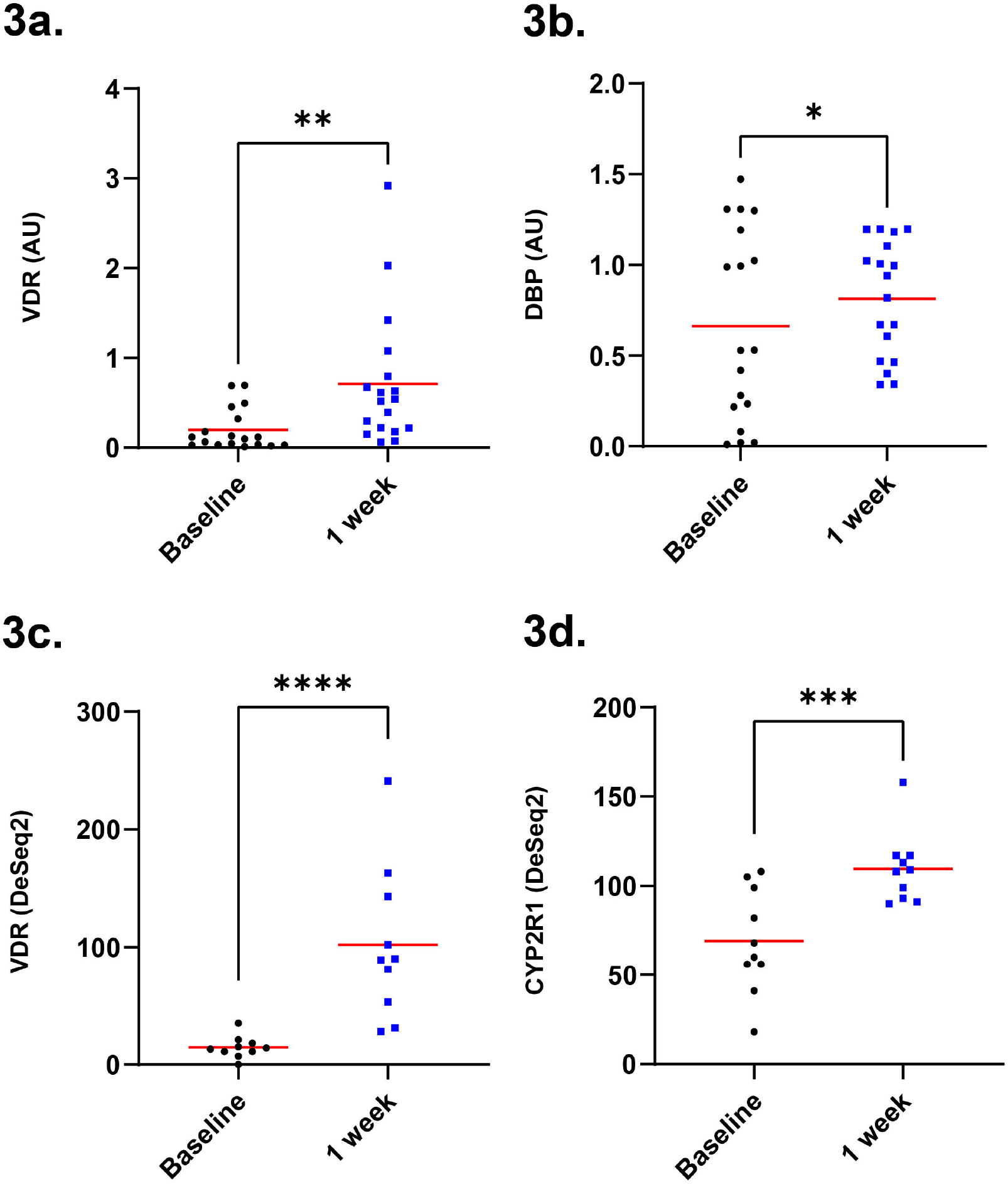
Vitamin D-associated transcripts and proteins in vastus lateralis elevated in response to ACL reconstruction (ACLR). 3a. Vitamin D receptor (VDR) protein arbitrary units (AU) on western blot increased in response to ACLR (n=18; p=0.006). 3b. Vitamin D binding protein (DBP) as indicated by western blots increased from baseline in response to ACLR (p=0.008). 3c. Vitamin D receptor (VDR) transcript count increased from baseline in response to ACLR (n=10 (post-ACLR)-12 (baseline)). 3d. Cytochrome P450 2R1 (CYP2R1) transcript count increased in response to ACLR. One-way repeated measures ANOVA; results of host-hoc tests on graph: * p-value<0.05; ** p-value <0.01; *** p-value<0.001. P-values presented in 3c and 3d reflect false discovery rate (FDR) adjustment for RNA-sequencing data.

### Low vitamin D status associates with CSA loss one week and four months after ACLR

Among participants with an average total 25(OH)D <30 ng/mL, CSA was lower 1 week and 4 months after ACLR (4455 ± 849 µm^2^ vs. 3285 ± 717 µm^2^ and 3119 ± 418 µm^2^, respectively; p<0.01 for post-hoc comparisons; p=0.041 for time x vitamin D status interaction), whereas those with total 25(OH)D ≥30ng/mL showed no significant differences (4291 ± 1046 µm^2^ vs. 4112 ± 1364 µm^2^ and 3867 ± 615 µm^2^, respectively; p>0.05 for all post-hoc comparisons). At the 4 month follow up, post-hoc analyses show CSA values were lower in the injured limb of participants having total 25(OH)D <30 ng/mL when compared with the 4-month CSA values of those having 25(OH)D ≥30ng/mL (3119 ± 418 µm^2^ vs. 3867 ± 615 µm^2^; p<0.01). CSA showed a trend toward lower values in the injured limb of participants having total 25(OH)D <30 ng/mL when compared with those having 25(OH)D ≥30ng/mL at 1 week post-ACLR (3285 ± 717 µm^2^ vs. 4112 ± 1364 µm^2^; p=0.051), and status groups showed no significant differences in CSA of the injured limb at baseline (4455 ± 849 µm^2^ vs. 4291 ± 1046 µm^2^; p=0.391). Fig. 4a shows CSA by vitamin D status group and complete data are presented in supplemental tables S3 and S4.

**Fig. 4.**
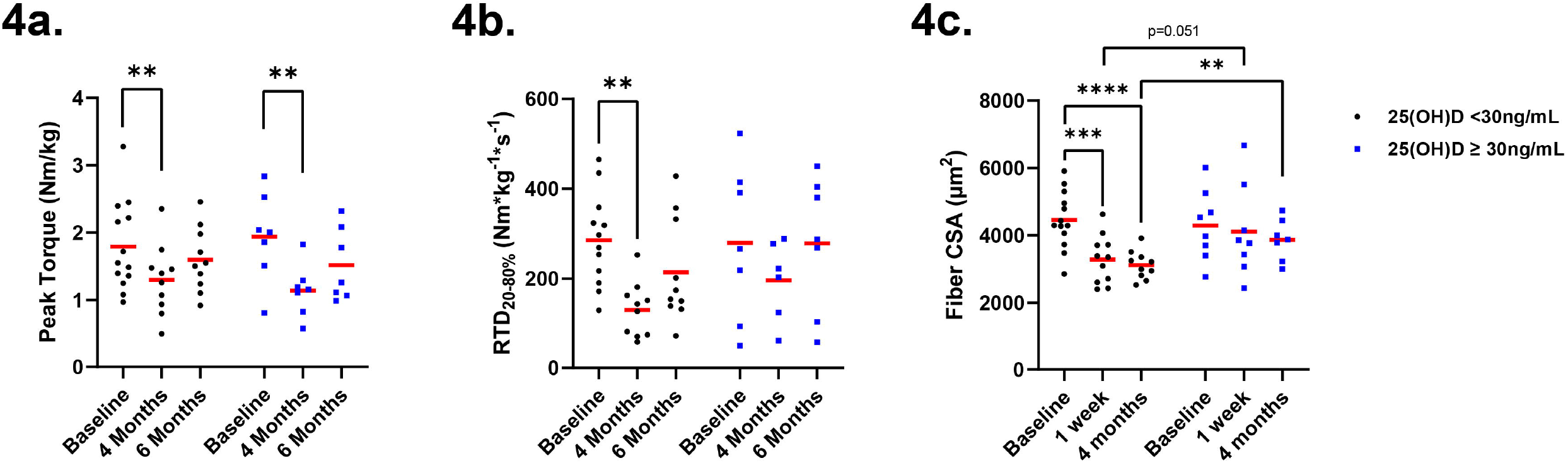
Mean study 25(OH)D <30ng/mL associates with cross sectional area (CSA) reductions in the vastus lateralis muscle but not strength or power. 4a. No differences in Maximum voluntary isometric contraction (MVIC) normalized peak torque in participants with study 25(OH)D <30ng/mL compared with participants having study 25(OH)D ≥30ng/mL. 4b. There is a significant loss of power over time but there is no differences between status groups. Both D status groups lose power as indicated by strength as determined by normalized maximal voluntary isometric contractions (peak torque). 4c. The low vitamin D status group shows significant reductions in fiber cross sectional area (CSA) at 1 week and 4 month follow ups when compared with baseline (n=13 in low status group). At 4 months, the fiber CSA is significantly lower in participants having study 25(OH)D <30ng/mL (n=13) when compared with participants who have 25(OH)D ≥30ng/mL (n= 8). Full-factorial repeated-measures ANOVA with post-hoc tests. The model showed an overall time by D status interaction effect p = 0.041; results of host-hoc tests on graph: * p-value <0.05, ** p-value <0.01, *** p-value <0.001, **** p-value <0.0001.

### Vitamin D status does not associate with strength, power or bone density outcomes

For all three BMD regions assessed, normalized peak torque, RTD_20-80%_, there was a significant main effect of time whereby all values decreased over time, on average. However, none of these indicators showed differences among participants with an average total 25(OH)D <30 ng/mL when compared with participants having a total 25(OH)D ≥30ng/mL (p>0.05 for status by time interactions). Data are presented in supplemental tables S5-8 and Fig. 4a, 4b, and Fig. 5.

**Fig. 5.**
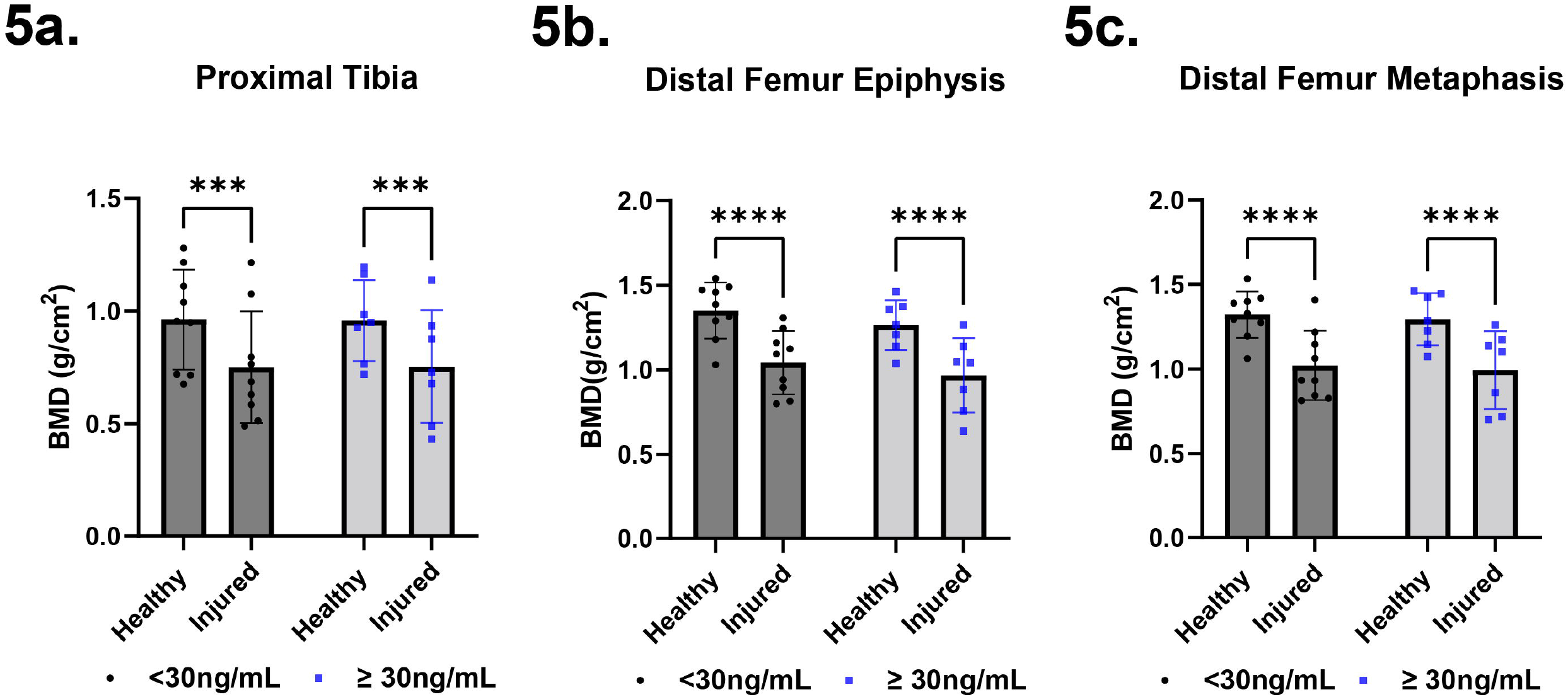
No differences in loss of bone mineral density (BMD) in participant with study 25(OH)D <30ng/mL compared with participants having study 25(OH)D <30ng/mL. 5a. BMD in proximal tibia. 5b. BMD in distal femoral epiphysis. 5c. BMD in distal femur metaphysis. Full-factorial repeated-measures ANOVA; results of host-hoc tests on graph: * p-value <0.05, ** p-value <0.01, *** p-value <0.001, **** p-value <0.0001.

## Discussion

Our results show that study participants with higher 25(OH)D concentrations (≥30ng/mL) were protected from quadriceps atrophy at 1 week and 4 months following ACL reconstruction. Additionally, we show that surgical reconstruction of the ACL induces expression of vitamin D receptors and enzymes within the quadriceps muscle acutely following surgery. These data provide intriguing evidence that vitamin D access and status are important nutritional considerations during and immediately after ACLR.

Prior studies have demonstrated that circulating vitamin D markers are negative acute phase reactants. In one study investigating patients receiving elective knee or hip surgery, both DBP and 25(OH)D were significantly reduced 2 days after surgery [37]. Others found that elective hip replacement surgery also promotes reductions in 1,25(OH)_2_D that are detectable several weeks after the procedure [38]. It is not clear whether our observed decreases in 1,25(OH)_2_D post-surgery, and significant increases in DBP at 4 and 6 month follow ups are due to decreased synthesis or increased catabolism or uptake, but our finding that DBP protein is elevated in skeletal muscle after ACLR indicates that the surgery may promote greater tissue uptake of vitamin D metabolites.

### Vitamin D status not associated with BMD loss

We did not observe any relationship between study vitamin D status defined with 25(OH)D and loss of BMD, and both groups experienced substantial loss of BMD in the three regions we assessed. Cross sectional studies have failed to show a strong association between circulating total 25(OH)D and BMD [39, 40]. Frank vitamin D deficiency clearly impairs bone mineralization [9], but marginal vitamin D status doesn’t seem to be a primary determinant of bone density loss in the injured leg following ACLR.

### Vitamin D status not associated with decrements in strength and power

Participants with 25(OH)D ≥30ng/mL did not show better maintenance of normalized peak torque or RTD_20-80%_ following ACLR when compared with those having concentrations <30ng/mL. Vitamin D supplementation studies in athletes generally have not demonstrated efficacy in strength or functional outcomes receiving placebo [41]. In one trial, adolescent swimmers having 25(OH)D <30ng/mL were instructed to consume vitamin D drops sufficient to provide 2000IU/day for 12 weeks with the goal of reaching 30ng/mL. Despite significant increases in circulating 25(OH)D and a 9.3 ng/mL difference between study groups at the trial’s conclusion, vitamin D supplemented participants did not increase grip strength or perform better on balance and swim tests when compared with the placebo group. Nonetheless, a meta-analysis of the effect of vitamin D supplementation on power, strength, and muscle mass showed small increases in muscle strength with vitamin D supplementation but no increases in power or muscle mass [17]. At the same time, older people with vitamin D concentration<12ng/mL show more substantial strength gains with vitamin D supplementation [17]. Gupta and others [42] showed that following ACLR, patients with 25(OH)D <20ng/mL had a graft failure rate of about 6% compared with a rate of 2% in patients with concentration of 30ng/mL; however, these outcomes were not statistically significant. Based on data from the Multicenter Orthopedic Outcomes Network (MOON) cohort, high body mass index, smoking, subsequent knee surgeries, and severe medial, lateral, and patellofemoral cartilage lesions are factors that predict functional outcomes 10 years after ALCR [43], but long-term relationship between diet and ACL outcomes have not been adequately evaluated.

### Clinical Implications

Vitamin D clinical cut points were established to prevent and treat frank vitamin D deficiency diseases like rickets and osteomalacia [9], but these same cut points may not be sufficient to optimize health and recovery from injury. Though there is not sufficient evidence here to conclude that increasing vitamin D concentrations to 30ng/mL will improve clinical outcomes in ACLR patients, this target is universally considered sufficient and not excessive. The Endocrine Society Clinical Practice Guidelines broadly supports an optimization cut point of 30ng/mL and indicates that children with concentrations of <20ng/mL may reach 30ng/mL by supplementing with 2000 IU/day (50 µg/day) for one year [44]. Adults may be supplemented with 50,000IU weekly (or 6000IU daily) for 8 weeks to reach 30 ng/mL. On a long term basis, vitamin D supplementation should not exceed the 4000 IU/day to avoid toxicity [9, 44]. These recommendations apply to healthy adolescents and adults and do not include people with aberrations in vitamin D or calcium metabolism.

### Limitations

Here our relatively small sample size did not allow us to separately analyze patients with very low circulating vitamin D, which has been more strongly associated with our outcomes [17]. The low sample and use of a single recruiting site also limits generalizability of results. It is possible that changes in vitamin D-associated markers may occur outside of the times that we collected samples. Though we were not powered to study modifying effects of skin tone, genetic background, or sex, all of these factors have the potential to modify vitamin D needs of athletes and should be studied intentionally moving forward.

### Conclusion

Our results demonstrate that vitamin D metabolism in whole skeletal muscle is elevated by ACLR. Associations between 25(OH)D<30 ng/mL and greater loss of CSA suggest that correcting vitamin D status prior to ACLR may support retention of skeletal muscle size in recovery, which should be tested in a randomized clinical trial to begin to establish vitamin D cut points optimizing recovery from ACL tear and reconstruction.

### Contributorship statement

Experiments were performed in the laboratory of JLF and CSF. BN, JLF and CSF acquired funding and were involved with conception and design of the experiments. JLF, ANM, CML, BML, NTT, and KAR collected and analyzed data. KLT and JLF performed study statistics. JLF drafted the manuscript and created figures. All authors revised and critically edited the manuscript for important intellectual content and approved the final version.

## Supporting information

Strobe Checklist

ICMJE J Fry

ICMJE A Moore

ICMJE C Latham

ICMJE K Thompson

ICMJE N Thomas

ICMJE B Lancaster

ICMJE C Fry

ICMJE K Reeves

ICMJE B Noehren

## Data Availability

All data produced in the present study are available upon reasonable request to the authors. RNA-sequencing data is publically available.

https://www.ncbi.nlm.nih.gov/geo/query/acc.cgi?acc=GSE211681

## Additional Information (including a Competing Interests Statement)

The authors do not declare any competing interests.

## Data Availability

The datasets generated during and/or analyzed during the current study are available from the corresponding author on reasonable request.

