## Supplement 1. Methods for "Vitamin D status associates with skeletal muscle loss after anterior cruciate ligament reconstruction"

*Supplemental Methods*

### ***Study Design***

The present manuscript uses data from an ongoing observational study where participants are enrolled prior to ACL reconstruction (ALCR). The aim of the parent study is to determine if acute induction of GDF-8 signaling following an ACL injury predicts reductions in muscle strength, connective tissue infiltration and dysregulation of skeletal muscle progenitor cells. For both the parent study and this manuscript, the contralateral limb serves as a control were relevant. Here, we executed this by using the contralateral limb as a covariate in all statistical assessments of clinical outcome variables. Summary of samples and data collected:

- Baseline: 1) Blood samples (serum separator tubes), strength testing, and DXA were completed within several days of study enrollment/consent. 2) Initial muscle biopsies were taken from the vastus lateralis of injured and contralateral limbs at the time of ACLR (most participants had ACLR within 7 days of study enrollment).
- One week post-ACLR: Blood and a muscle biopsy from the injured limb were collected
- One and two months: Blood was collected. We did not analyze any of these samples because these time points didn’t align with any of the outcomes, e.g. skeletal muscle fiber cross sectional area (CSA), bone mineral density (BMD), or Biodex.
- 4 months: Blood, Biodex data, and the final muscle biopsy from the injured limb were collected.
- 6 months: Blood, Biodex and the final BMD data were collected.

CSA, BMD, Biodex, and RNA-sequencing analyses were collected in the original parent study. All circulating markers and western blots were completed for this manuscript as part of a separate training grant. All participants having completed baseline and at least one follow up session at the conclusion of the training grant were included in the analysis for this manuscript. Thirty participants signed the consent form, seven participants withdrew before ACLR, and two were participants withdrew after data collection 1 week after surgery. The analysis includes 21 participants and 19 of these completed the 6 month study. All available samples were used to generate data for analysis and values of missing data points were not imputed.

### ***Location and Recruitment***

Bone-patellar-bone graft ACL surgeries were completed by a single physician at the University of Kentucky Orthopedic Surgery & Sports Medicine practice in Lexington, KY (latitude of 38.0406° N). The physicians in the practice complete 200+ ACL reconstruction surgeries each year. A clinical coordinator embedded in the clinic was responsible for recruiting subjects and monitoring their progress throughout the study. All study protocols were approved by the University of Kentucky Institutional Review Board. All participants were provided written and oral consent prior to data collection or parental consent and child assent, where applicable.

***Participants***

All healthy participants meeting the inclusion criteria were invited to participate. All participants (n=21) were recruited after an ACL injury and before ACLR surgery, were between 15-45 years of age, underwent bone-patellar-bone graft ACLR conducted by the same surgeon (no prophylactic antibiotics), and completed a progressive rehabilitation program according to previously published guidelines at the University of Kentucky’s Physical Therapy Department. Patients with total knee dislocation were excluded but those with meniscus tear remained eligible.

#### *Circulating Biomarkers*

Serum samples were collected prior to ACLR, 1-week post ACLR, and at 4 and 6 month follow ups. Study vitamin D status was determined by averaging 25-hydroxy vitamin D (25(OH)D) over 4 time points (i.e. baseline and 1 week, 4 month, and 6 month follow ups). We sent 0.5mL serum samples to Mayo Clinic Laboratories to assess 25(OH)D using gold standard LC-MS/MS methodology. ELISA was used to assess 1,25-dihydroxy vitamin D (1,25(OH)2D; Biovendor R&D RIS024R and RIS021R), free 25(OH)D (Biovendor R&D KAPF1991) and vitamin D binding protein (DBP; R&D Systems DY008B and DY3778B-05) according to the manufacturer’s instructions. All commercially available DBP assays require substantial dilution of the serum sample to get the analyte in the range of the assay. Human DBP is approximately 200-300µg/mL, and the assay we used has a range of 187.0 - 3,000 pg/mL. We diluted all samples 1:500,000 according to the table below. We repeated a plate (about ½ of samples).

| Tube # | Sample to Dilute | Volume of Sample (µL) | Volume of 1X Wash Buffer (µL) | Volume of Sample Diluent NS (µL) | Starting Conc. | Final Conc. |
| --- | --- | --- | --- | --- | --- | --- |
| 1 | Neat Serum | 5 | 995 | - | Neat | 1:200 |
| 2 |  | 5 | 495 | - | 1:1000 | 1:20,000 |
| 3 |  | 5 | - | 120 | 1:20,000 | 1:500,000 |

We repeated a plate (containing about ½ of samples used) diluting 1:405,224. Our average interassay CV was 10.5%. For all circulating markers assessed with ELISA, all samples for each participant were run on the same plate. Results were read on a SpectraMax M2 plate reader.

*Muscle Biopsies*

Biopsies were taken from the vastus lateralis on the injured and contralateral healthy limbs at the time of ACLR and from the injured limb only 1 week and 4 months after ACLR. Participants received local anesthetic (1% Xylocaine HCl) and biopsies were collected using a scalpel during ACLR and at follow up visits with modification of Bergstrom's percutaneous biopsy technique which provides for 100mg of muscle tissue for immunohistochemistry (IHC) analysis and protein/gene expression analyses. The sample was divided and flash frozen for RNA/protein and for IHC mounted in tragacanth using a dissecting microscope and frozen in isopentane cooled to the temperature of liquid nitrogen.

#### *Western Blot*

Biopsies were homogenized using a buffer consisting of 50 mM Tris-HCL, 250 mM mannitol, 50 mM NaF, 5 mM Na pyrophophate, 1 mM EDTA, 1 mM EGTA. For each 50mL of complete buffer, 7.5mg Dithiothreitol (DTT) 99%, 7.8 benzamidine hydrochloride hydrate 98%, 25 µL 200mM PMSF, and 250 µL HALT protease/phosphatase inhibitor cocktail (Thermo Fisher 87786) were added immediately before use. Approximately 30mg of each muscle sample was thoroughly homogenized using a bullet blender tissue homogenizer (Next Advance, Inc., Troy, NY) in a 4°C room using 9 µL of buffer per mg of tissue. Homogenate was transferred to a microcentrifuge tube and centrifuged for 10 minutes at 6000rpm at 4°C. The supernatant was collected and total protein was determined with the Bradford assay (Smartspec Plus spectrophotometer; Bio-Rad, Hercules, CA, USA). Supernatant was combined with equal parts sample buffer (2x Laemmli Sample Buffer #1610737 (Bio-Rad) with 2-Mercaptethanol). Kidney lysate (VDR positive control) and HELA lysate (loading control) were run on all gels using the same methods. Samples containing 50µg of protein per well were loaded on to a 4–20% Criterion™ TGX Stain-Free™ Protein 18 well gel (Bio-rad) and run at 150 volts for 80 minutes in Tris/Tricine/SDS Running Buffer (Bio-Rad #1610744). Stain-free technology was activated with the Chemidoc Touch imager (Bio-Rad). Protein was transferred to a methanol-actviated Immun-Blot LF PVDF membrane (Bio-Rad) for 50 volts for 60 minutes in transfer buffer (10 mM 3-(cyclohexylamino) propane-1-sulfonic acid, pH 11, 10% methanol). Total protein was imaged on the Chemidoc before adding primary antibodies. After washing in TBST, membranes were blocked in 5% bovine serum albumin (BSA) in TBST for one hour. Blots were incubated in primary antibodies overnight at room temperature, washed, incubated in an appropriate HRP-conjugated secondary antibody, incubated with ECL (Thermo Scientific 80196X3) for 5 minutes, and imaged in the chemiluminescent channel on the Chemidoc. Blots were probed with VDR antibodies (Abcam ab109234; 1:1000 in 5% Bovine Serum Albumin in TBST), then stripped and blocked before incubating overnight in DBP antibodies (0.25µg/mL; R&D Systems DY3778B-05 detection antibody). All blots were analyzed in ImageLab (Bio-Rad) by creating a multi-channel image with total protein coupled to the chemiluminescent channel to normalize for total protein loaded into wells. For each participant, all of their samples were loaded on the same gel.

#### *RNA isolation and sequencing (RNA-seq)*

Quadriceps muscle tissue (~25-30mg) was homogenized in Tri-reagent using Zirconium Oxide beads (2.0 mm, RNase-free, Next Advance, Inc. Troy, NY). The homogenate was used to isolate total RNA in accordance with manufacturer guidelines (Direct-zol RNA Miniprep Kit, Zymo) 1. RNA content, purity, and integrity was quantified using the 2100 Bioanalyzer (Agilent) (RIN > 8.5) and the NanoDrop 2000 (Thermo Fisher) at the University of Kentucky Genomics Core. Six hundred nanograms of total RNA was sent to Novogene Corporation (Chula Vista, CA) for library construction and sequencing on an Illumina HiSeq 4000 system using a paired-end 150 bp dual-indexing protocol. Raw FASTQ files underwent pre-alignment quality control, then were aligned using STAR aligner (reference genome: GRCh38/hg38) using Partek Flow, and analyzed to obtain differential gene expression using DESeq2 (minimum read cutoff of 50). Differential gene expression was calculated comparing ACL-injured samples collected during surgery and 1 week post-ACLR and comparing healthy limb samples collected during surgery and 1 week post-ACLR. Raw p-values were adjusted for multiple testing using the Benjamini-Hochberg false discovery rate (FDR) step-up method. Pathway over-representation analysis was performed using g:Profiler with non-ordered query and up-or-downregulated genes with FDR < 0.05. RNA-sequencing data are deposited in Gene Expression Omnibus: GSE211681. Transcript abundance of vitamin D activating pathway regulators (vitamin D receptor [*VDR*], and Cytochrome P450 2R1 [*CYP2R1*] and Family 27 Subfamily B Member 1 [*CYP27B1*]) are reported here.

#### *Immunohistochemistry Analyses*

Cross-sectional area was determined by cryosectioning 7µm thick sections using the HM525X cryostat (ThermoFisher, Waltham, MA) and mounting sections on charged slides. Sections were air dried at room temperature overnight, then rehydrated in PBS for 3 minutes to equilibrate sections to PBS. Following rehydration, slides were incubated in a rabbit-raised primary antibody against laminin (Sigma, cat# L9393) (diluted 1:100 in PBS) overnight. Slides then underwent 3 serial washes in PBS for 3 minutes each. Sections were then incubated in AlexaFluor 555 goat anti-rabbit secondary antibody (Invitrogen, cat# A21429) (diluted 1:250 in PBS) for 2 hours, mounted with Vectashield mounting media (Vector Laboratories, cat# H-1000) and imaged at 10x (100x total magnification) on a Ziess AxioImager M2 upright fluorescent microscope.

We used established, automated ICH protocols for assessing average muscle fiber cross sectional area (CSA) using Myovision as previously described [31]. For image data acquisition, MyoVision, an automated image analysis application, was used 1. Raw (.CZI) image files were exported as 24 bit .PNG files with no compression prior to running the images through MyoVision to automatically assess fiber CSA.

#### *Strength Outcomes*

Participants’ maximum voluntary isometric contraction (MVIC) and the mean slope of the torque-time curve between 20% and 80% of the first 200 milliseconds from muscle contraction onset (RTD20-80%) were evaluated before ACLR and at 4 and 6 month follow ups as previously reported (Kline 2015). Muscle contraction onset was defined as the point when the torque signal surpassed a 6 Nm threshold. Participants completed MVIC and RTD20-80% testing in both limbs using a Biodex 4 isokinetic dynamometer (Biodex Medical Systems Inc., Shirley, NY, USA) with the subject’s knee and hip placed at 90 degrees of flexion and secured with straps to limit extraneous movements. The dynamometer attachment arm was secured to the shank approximately 5 cm proximal to the medial malleolus. After practicing once, participants completed four subsequent test trials and were instructed to kick and hold as hard and as quickly as they could for 5 seconds while strong verbal encouragement was given. Participants rested for 30 seconds between sets and 5 minutes rest between limbs. The torque signal was sampled at 100 Hz and filtered using a fourth-order, low-pass Butterworth zero-lag digital filter with a 24 Hz cut-off frequency. Results were analyzed with custom MATLAB code as previously described (Kline 2015).

*Bone Density Measures*

Bone mineral density (BMD) was assessed with DXA scans (Lunar iDXA, GE Healthcare) and were completed at study baseline and at the 6 month follow up in both the injured and healthy limbs. We
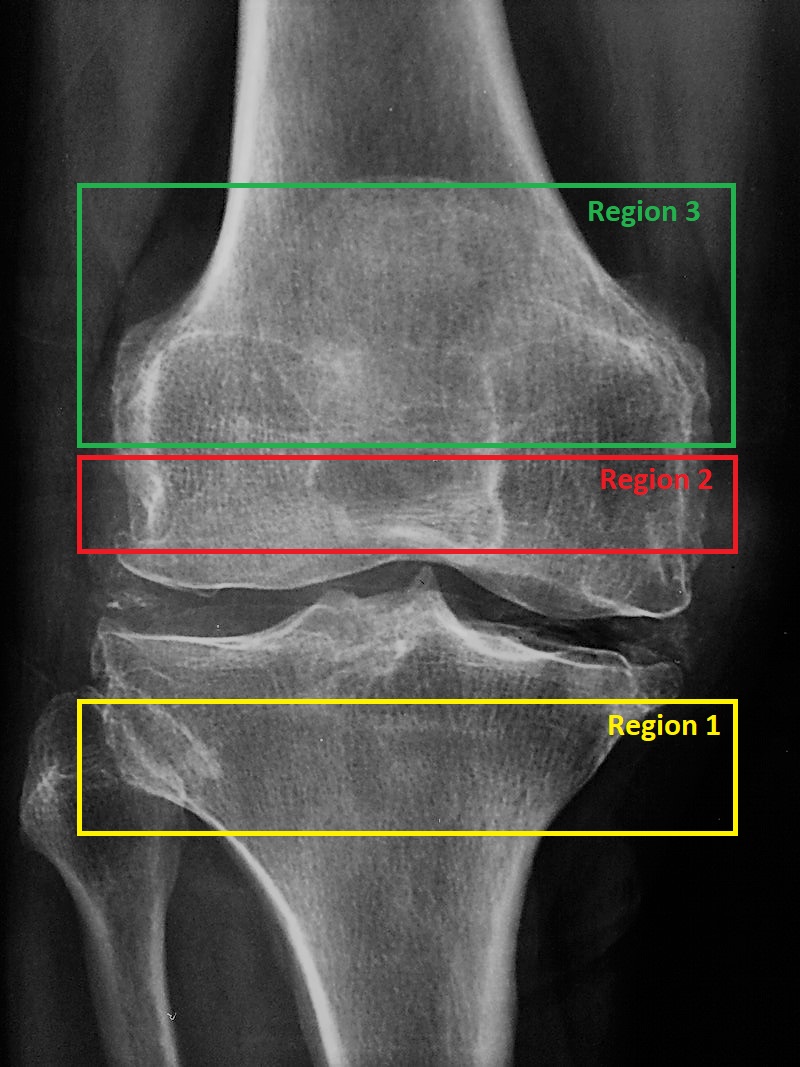
utilized a validated protocol for determining BMD in the femur and tibia. Participants completed the DXA scan lying on their backs while fully extending the scanned leg with the supported and foot rotated toward midline. Participants were instructed to remain still and all scans clearly depicted a horizontal joint space, the proximal patella and distal fibular head. For consistency between scans over the time course, the DXA laser light was positioned at 3.5 cm above the proximal border of the patella. Three regions were measured: Region 1- the proximal tibia was measured by the distal horizontal edge of the region of interest placed at the most distal point of contact between the fibular head and the tibia. The proximal edge was placed at the upper edge of the fibular head; Region 2- the most distal aspect of the femur was measured with the bottom horizontal edge of the region of interest positioned at the top of the joint line between the femoral condyles with a height set to match that of the proximal tibia; and Region 3- the distal femur encompassing the whole patella was measured with the exact bottom horizontal edge as in region 2, with the proximal edge set higher than region 2 positioned at the upper edge of the patella. The DXA enCORE software platform automatically calculated bone mineral density.

Figure 1. Knee MRI image showing analysis regions. Image obtained with a CREATIVE commons license from https://www.rawpixel.com/image/3306165/free-photo-image-medical-hospital

*Statistical Analyses*

For all outcomes, statistical significance was set at *p* < .05, using two-sided tests and using adjusted *p*-values where appropriate. All continuous measures were summarized with descriptive statistics, and distributions were tested for normality. Circulating measures assessed at 4 time points were analyzed using a mixed-effects repeated measures analysis using the Tukey post-hoc test to adjust for multiple comparisons (n=21). Results of western blots were analyzed using paired T-tests from the injured limb at baseline and the injured limb one week after surgery (n=18). To assess relationships between vitamin D status and CSA, 25(OH)D concentrations from all time points were averaged prior to defining status as dichotomous variable based on the a priori optimization cut-off value of ≥30 ng/mL to determine low and high status groups with those having a study mean 25(OH)D ≥ 30ng/mL included in the high group (n=8) and those with concentrations < 30ng/mL included in the low group (n=13). Analyzing vitamin D status groups as a dichotomous variable with a cut point is 30ng/mL is common [13, 35, 36]. Since CSA, DXA, and Biodex measures included multiple observations taken from the same subject over the injured/non-injured legs and across multiple visits, a full-factorial repeated-measures ANOVA was performed, first analyzing overall differences across the various treatment groups (time point/leg and vitamin D status). Likelihood ratio testing and Akaike Information Criterion (AIC) were used to select an appropriate covariance structure (here, compound symmetry covariance). A Kenward-Roger adjustment was used, as appropriate, to correct for negative bias in the standard errors and degrees of freedom calculations induced by small samples. For each relevant pairwise comparison, estimated differences of means (calculated as Group 1 (adequate status) - Group 2 (low status)) and the associated standard errors are adjusted for baseline (V2) non-injured leg value. Efforts were made to align with the BJSM checklist for statistical assessment of medical papers and justify deviations were relevant [34]. All analyses were completed in SAS 9.4 (SAS Institute Inc.; Cary, NC, USA).
