## Supplement 2. Tables for "Vitamin D status associates with skeletal muscle loss after anterior cruciate ligament reconstruction"

Supplementary Table S1.Circulating Vitamin D Indicators.

|  | Baseline  (n=21) | 1-wk Post Surgery  (n=20) | 4 months  (n=17) | 6 months  (n=16) | p-value  (main time effect) | p-value  (post-hoc) |
| --- | --- | --- | --- | --- | --- | --- |
| 25(OH)D^1^ (ng/mL) | 30.1 ± 12.4 | 27.1 ± 10.5 | 28.5 ± 9.2 | 31.6 ± 12.4 | 0.474 | N/A |
| 1,25(OH)_2_D^2^ (pg/mL) | 21.6 ± 7.9 | 13.8 ± 5.5 | 23.5 ± 13.4 | 23.4 ± 6.1 | 0.003 | BL^5^ vs. 1-wk: <0.001  1wk^6^ vs 4mo: 0.003  1wk. vs. 6 mo: 0.003 |
| Free 25(OH)D^3^ (pg/mL) | 7.0 ± 3.2 | 6.1 ± 2.3 | 6.1 ± 2.7 | 6.8 ± 2.5 | 0.062 | N/A |
| DBP^4^ (µg/mL) | 233.2 ± 103.4 | 221.9 ± 60.6 | 200.5 ± 99.2 | 192 ± 73.6 | 0.0009 | BL vs 4mo: 0.016  BL vs. 6 mo: 0.027 |

^1^ 25-hydroxyvitamin D

^2^ 1,25-dihydroxyvitamin D

^3^ Free 25-hydroxyvitamin D (unbound to protein carrier)

^4^ Vitamin D binding protein

^5^ Baseline visit before anterior cruciate ligament reconstruction (ACLR)

^6^ One week after ACLR

Supplementary Table S2.Circulating Vitamin D Indicators by Study 25(OH)D Status Group.

|  | Baseline | | 1-wk Post Surgery | | 4 months | | 6 months | |
| --- | --- | --- | --- | --- | --- | --- | --- | --- |
|  | Low  (n=13) | High  (n=8) | Low  (n=13) | High  (n=7) | Low  (n=10) | High  (n=7) | Low  (n=9) | High  (n=7) |
| 25(OH)D^1^ (ng/mL) | 22.5 ± 5.3 | 42.4 ± 10.6 | 21.2 ± 6.3 | 38±7.5 | 24.5 ± 8.4 | 34.3 ± 7.3 | 25.5 ± 7.6 | 41.2 ± 12.5 |
| 1,25(OH)_2_D^2^ (pg/mL) | 19.7 ± 6.9 | 24.6 ± 8.9 | 13.4 ± 4.5 | 14.6 ± 7 | 19.5 ± 7 | 29.2 ± 18.4 | 23.2 ± 4.9 | 24 ± 7.6 |
| Free 25(OH)D^3^ (pg/mL) | 5.1 ± 1.6 | 9.2 ± 2.3 | 4.7 ± 1.4 | 8.2 ± 1.3 | 5.7 ± 2.7 | 7.7 ± 2.4 | 5.7 ± 2.1 | 8.4 ± 2.0 |
| DBP^4^ (µg/mL) | 227.4 ± 57.1 | 242.6 ± 157.5 | 235.6 ± 53.3 | 196.3 ± 69.1 | 185.2 ± 49.6 | 222.3 ± 147 | 188.4 ± 44.5 | 186.7 ± 104.2 |

^1^ 25-hydroxyvitamin D

^2^ 1,25-dihydroxyvitamin D

^3^ Free 25-hydroxyvitamin D (unbound to protein carrier)

^4^ Vitamin D binding protein

Supplementary Table S3. Vitamin D Expresssion Markers.

|  | Baseline | | 1 Week After Surgery  Injured | 4 Months After Surgery  Injured | p-value  (main time effect) |
| --- | --- | --- | --- | --- | --- |
|  | Non-Injured | Injured |  |  |  |
| DBP^1^ (AU; n=18) | 0.55 ± 0.48 | 0.66 ± 0.53 | 0.81 ± 0.32 | - | 0.047 |
| VDR^2^ (AU; n=18) | 0.4 ± 0.5 | 0.2 ± 0.2 | 0.7 ± 0.7 | - | 0.008 |
| Fiber CSA^3^ (µm^2^; n=21) | 4777 ± 1006 | 4394 ± 906 | 3633 ± 1088 | 3428 ± 620 | 0.043 |
| *VDR* RNA-seq (n=10-12) | 11 ± 9 | 13 ± 9 | 102 ± 65 | - | <0.001 |
| *CYP2R1*^4^ RNA-seq | 55 ± 22 | 67 ± 29 | 110 ± 20 | - | 0.0009 |
| *CYP27B1*^5^ RNA-seq | 3 ± 4 | 3 ± 3 | 7 ± 5 | - | 0.1288 |

^1^ Vitamin D binding protein in quadriceps sample

^2^ Vitamin D receptor protein in quadriceps sample

^3^ Quadriceps muscle fiber cross sectional area

^4^ Cytochrome P450 Family 2 Subfamily R Member 1 (i.e. 25-hydroxylase)

^5^ Cytochrome P450 Family B1 Subfamily B Member 1 (i.e. 1α-hydroxylase)

Supplementary Table S4. Vitamin D Expresssion Markers by D status Group

|  | Baseline | | | | 1 Week After Surgery  (Injured Only) | | 4 Months After Surgery  (Injured Only) | |
| --- | --- | --- | --- | --- | --- | --- | --- | --- |
|  | Non-Injured | | Injured | |  |  |  |  |
|  | Low | High | Low | High | Low | High | Low | High |
| DBP^1^ (AU; n=18) | 0.47 ± 0.46 | 0.66 ± 0.51 | 0.59 ± 0.47 | 0.78 ± 0.63 | 0.78 ± 0.32 | 0.86 ± 0.34 | - | |
| VDR^2^ (AU; n=18) | 0.2 ± 0.3 | 0.6 ± 0.6 | 0.2 ± 0.2 | 0.2 ± 0.2 | 0.8 ± 0.9 | 0.6 ± 0.4 | - | |
| Fiber CSA^3^ (µm^2^; n=21) | 4630 ± 886 | 5016 ± 1200 | 4454 ± 849 | 4292 ± 1046 | 3285 ± 717 | 4112 ± 1364 | 3120 ±418 | 3867 ± 615 |
| *VDR* RNA-seq (n=10) | 9 ± 10 | 20 ± 2 | 14 ± 9 | 11 ± 8 | 110 ± 64 | 29 ± 0 | - | |
| *CYP2R1*^4^ RNA-seq (n=10) | 60 ± 15 | 37 ± 34 | 72 ± 29 | 44 ± 18 | 110 ± 21 | 108 ± 0 | - | |
| *CYP27B1*^5^ RNA-seq (n=10) | 3 ± 4 | 3 ± 3 | 3 ± 3 | 4 ± 1 | 7 ± 5 | 5 ± 0 | - | |

^1^ Vitamin D binding protein in quadriceps sample

^2^ Vitamin D receptor protein in quadriceps sample

^3^ Quadriceps muscle fiber cross sectional area

^4^ Cytochrome P450 Family 2 Subfamily R Member 1 (i.e. 25-hydroxylase)

^5^ Cytochrome P450 Family B1 Subfamily B Member 1 (i.e. 1α-hydroxylase)

Supplementary Table S5. Dual Energy X-Ray Absorptiometry Bone Mineral Density (BMD)

|  | Baseline  (n=18) | | 6-months after ACLR^1^  (n=16) | | p-value  (main time effect) |
| --- | --- | --- | --- | --- | --- |
| BMD Region | Non-injured | Injured | Non-injured | Injured |  |
| Proximal Tibial Epiphysis (g/cm^2^) | 1.00 ± 0.20 | 0.96 ± 0.21 | 0.96 ± 0.20 | 0.75 ± 0.24 | 0.002 |
| Distal Femoral Epiphysis (g/cm^2^) | 1.32 ± 0.17 | 1.30 ± 0.18 | 1.31 ± 0.16 | 1.01 ± 0.20 | <0.001 |
| Distal Femoral Metaphysis (g/cm^2^) | 1.3 ± 0.16 | 1.27 ± 0.17 | 1.31 ± 0.14 | 1.01 ± 0.21 | <0.001 |

^1^ Anterior cruciate ligament reconstruction

Supplementary Table S6. Dual Energy X-Ray Absorptiometry Bone Mineral Density (BMD) by D Status Group

|  | Baseline | | | | 6 Months after ACLR | | | |
| --- | --- | --- | --- | --- | --- | --- | --- | --- |
|  | Non-Injured | | Injured | | Non-Injured | | Injured | |
| BMD Region | Low^1^  (n= 12) | High^2^  (n=6 ) | Low  (n= 12) | High  (n=6 ) | Low  (n= 9) | High  (n=7 ) | Low  (n= 9) | High  (n=7 ) |
| Proximal Tibial Epiphysis (g/cm^2^) | 1.00 ± 0.20 | 1.01 ± 0.23 | 0.95 ± 0.15 | 0.97 ± 0.31 | 0.96 ± 0.22 | 0.96 ± 0.18 | 0.75 ± 0.25 | 0.75 ± 0.25 |
| Distal Femoral Epiphysis (g/cm^2)^ | 1.34 ± 0.17 | 1.29 ± 0.18 | 1.31 ± 0.16 | 1.29 ± 0.22 | 1.35 ± 0.17 | 1.26 ± 0.15 | 1.04 ± 0.19 | 0.97 ± 0.22 |
| Distal Femoral Metaphysis (g/cm^2^) | 1.34 ± 0.14 | 1.31 ± 0.20 | 1.28 ± 0.12 | 1.26 ± 0.25 | 1.32 ± 0.138 | 1.29 ± 0.15 | 1.02 ± 0.20 | 0.99 ± 0.23 |

Supplementary Table S7. Strength and Power Indicators.

|  | Baseline  (n= 19-21) | | | 4 Months after ACLR^1^  (n= 16-17) | | | 6 Months after ACLR  (n= 16-17) | | |
| --- | --- | --- | --- | --- | --- | --- | --- | --- | --- |
|  | Non-injured | Injured | LSI | Non-injured | Injured | LSI | Non-injured | Injured | LSI |
| Normalized Peak Torque (Nm/kg) | 2.43± 0.65 | 1.85 ± 0.65 | 0.77 ± 0.23 | 2.73 ± 0.66 | 1.24 ± 0.47 | 0.47 ± 0.13 | 1.323 ± 0.169 | 1.57 ± 0.49 | 0.60 ± 0.14 |
| RTD^2^_20-80%_ (Nm*kg-1*s-1)) | 406.6 ± 202.7 | 283.6 ± 128.4 | 0.72 ± 0.26 | 476.2 ± 2511.3 | 154.9 ± 77.3 | 0.38 ± 0.18 | 501.6 ± 269.4 | 240.8 ± 131.1 | 0.54 ± 0.33 |

^1^ Anterior cruciate ligament reconstruction

^2^ Peak torque as indicated by maximum voluntary isometric contraction normalized to body weight

^3^ Rate of torque development from the mean slope of the torque-time curve between 20% and 80% of the first 200 milliseconds from muscle contraction onset

Supplementary Table S8. Strength and Power Indicators by D Status Group

|  | **Baseline** | | | | **4 Months after ACLR^1^** | | | |
| --- | --- | --- | --- | --- | --- | --- | --- | --- |
|  | Non-Injured | | Injured | | Non-Injured | | Injured | |
|  | Low  (n=13) | High  (n=8) | Low  (n=13) | High  (n=8) | Low  (n=10) | High  (n=7) | Low  (n=10) | High  (n=7) |
| Normalized Peak Torque (Nm/kg)^2^ | 2.44 ± 0.61 | 2.37 ± 0.72 | 1.79 ± 0.67 | 1.76 ± 0.80 | 2.74 ± 0.63 | 2.71 ± 0.74 | 1.30 ± 0.52 | 1.14 ± 0.39 |
| RTD^3^_20-80%_ (Nm/kg^-1^*s^-1^) | 439.3 ± 214.3 | 345.8 ± 177.9 | 286.0 ± 102.2 | 279.5 ± 173.9 | 480.1 ± 275.9 | 469.6 ± 228.0 | 130.1 ± 61.4 | 196.3 ± 88.7 |
|  |  |  |  |  | **6 Months after ACLR** | | | |
|  |  |  |  |  | Non-Injured | | Injured | |
|  |  |  |  |  | Low  (n=10) | High  (n=6-7) | Low  (n=10) | High  (n=6-7 ) |
|  |  |  | Normalized Peak Torque (Nm/kg) | | 2.73 ± 0.32 | 2.51 ± 0.83 | 1.60 ± 0.47 | 1.52 ± 0.54 |
|  |  |  | RTD_20-80%_ (Nm/kg^-1^*s^-1^) | | 503.4 ± 273.6 | 498.8 ± 288.0 | 214.2 ± 116.6 | 278.7 ± 150.3 |

^1^ Anterior cruciate ligament reconstruction

^2^ Peak torque as indicated by maximum voluntary isometric contraction normalized to body weight

^3^ Rate of torque development from the mean slope of the torque-time curve between 20% and 80% of the first 200 milliseconds from muscle contraction onset
